# Semantic speech networks linked to formal thought disorder in early psychosis

**DOI:** 10.1101/2022.02.25.22271517

**Authors:** Caroline R Nettekoven, Kelly Diederen, Oscar Giles, Helen Duncan, Iain Stenson, Julianna Olah, Toni Gibbs-Dean, Nigel Collier, Petra E Vértes, Tom J Spencer, Sarah E Morgan, Philip McGuire

## Abstract

**Background and Hypothesis:** Mapping a patient’s speech as a network has proved to be a useful way of understanding formal thought disorder in psychosis. However, to date, graph theory tools have not incorporated the semantic content of speech, which is altered in psychosis.

**Study Design:** We developed an algorithm, “*netts*”, to map the semantic content of speech as a network, then applied *netts* to construct semantic speech networks for a general population sample, and a clinical sample comprising patients with first episode psychosis (FEP), people at clinical high risk of psychosis (CHR-P), and healthy controls.

**Study Results:** Semantic speech networks from the general population were more connected than size-matched randomised networks, with fewer and larger connected components, reflecting the non-random nature of speech. Networks from FEP patients were smaller than from healthy participants, for a picture description task but not a story recall task. For the former task, FEP networks were also more fragmented than those from controls; showing more, smaller connected components. CHR-P networks showed fragmentation values in-between FEP patients and controls. A clustering analysis suggested that semantic speech networks captured novel signal not already described by existing NLP measures. Network features were also related to negative symptom scores and scores on the Thought and Language Index, although these relationships did not survive correcting for multiple comparisons.

**Conclusions:** Overall, these data suggest that semantic networks can enable deeper phenotyping of formal thought disorder in psychosis. We are releasing *Netts* as an open Python package alongside this manuscript.

## Introduction

One of the great challenges facing psychiatry is mapping the complexity and heterogeneity of mental health conditions, including psychotic disorders. Although psychotic disorders are recognized for their multifaceted and diverse phenotypes, objectively describing and measuring these remains difficult. Generating new approaches for the automated analysis and quantification of psychotic symptoms could not only significantly advance our understanding of this debilitating condition, but also aid disease monitoring and prediction.

A core symptom of psychosis is formal thought disorder (FTD), manifesting as changes in the patient’s speech which can appear incoherent and disorganized. FTD has several dimensions, as described by the Thought and Language Index (1), including poverty of speech and loosening of associations, where the connection between ideas is tenuous or extraneous ideas intrude into the train of thought (1). Several studies have highlighted the importance of FTD. A large longitudinal study showed loosening of associations in speech appeared early in the disorder, was present in the majority of cases and did not resolve over thirty years of investigation (2). More generally, FTD has been related to more severe forms of psychosis (3, 4), lower quality of life (5) and has been suggested to predict functional outcomes and relapse (6).

The advent of natural language processing (NLP) techniques has provided new computational approaches to measure disorganised speech and FTD (7–13). These approaches can be applied automatically by computers and are therefore substantially less time-consuming than manual assessments using the TLI. For example, (13) created networks to measure disorganised speech, in which nodes were represented as words, and edges connected words in the order in which they were spoken. These word-trajectory networks showed patient-control differences in speech and predicted diagnosis 6 months in advance (11).

However, these networks focus on the syntactic characteristics of speech and largely ignore semantic content. Meanwhile, measuring semantic coherence has been found to predict psychotic disorders (8, 14), but only provides a single summary measure of an entire speech transcript. Speech excerpts likely include additional semantic information that could be used to build a deeper understanding of how the semantic content of language is altered in schizophrenia; and provide extra power to predict outcome or relapse for individual patients. Networks provide a natural way to represent the semantic content of a speech transcript in more detail, building on the idea that “reality is knowable as a set of informational units and relations among them” (15, 16). Hence it is plausible that representing the semantic content of speech as a network could shed fresh light on the nature of speech in psychosis.

We therefore developed a novel speech network algorithm that maps the semantic content of transcribed speech, creating *semantic speech networks*. These semantic speech networks provide a natural framework to capture the information conveyed by the speaker by representing the entities (e.g. a person, object or colour) the speaker mentions as nodes and the relationships between entities as edges in the network. We can then test whether the properties of these networks and relationships are related to particular properties of FTD, such as speech fragmentation and loosening of associations. We are releasing the tool as an openly available Python package named **Networks of Transcribed Speech in Python (*netts*)** alongside this paper. The tool can be installed from PyPI (17) and used to construct a semantic speech network from a text file with a single command.

In the following, we introduce *netts* and outline the processing steps the algorithm takes to construct a semantic speech network from transcribed speech. We then describe results from applying *netts* to speech transcripts from a general population sample, where we explored the general properties of semantic speech networks and compared them to random networks. We also used the tool to test for group differences between semantic speech networks of first episode psychosis patients, individuals at clinical high risk for psychosis and healthy controls. Finally, we explore the relationships of the semantic speech network measures with symptom severity, manual Thought and Language Index scores and with other established NLP markers.

## Methods

### Network Algorithm

To construct semantic speech networks we created *netts. Netts* takes as input a speech transcript (e.g. *I see a man*) and outputs a semantic speech network, where nodes are entities (e.g. *I, man*) and edges are relations between nodes (e.g. *see*).

In the following we describe the *netts* pipeline, illustrated in Figure 1.

**Fig. 1.**
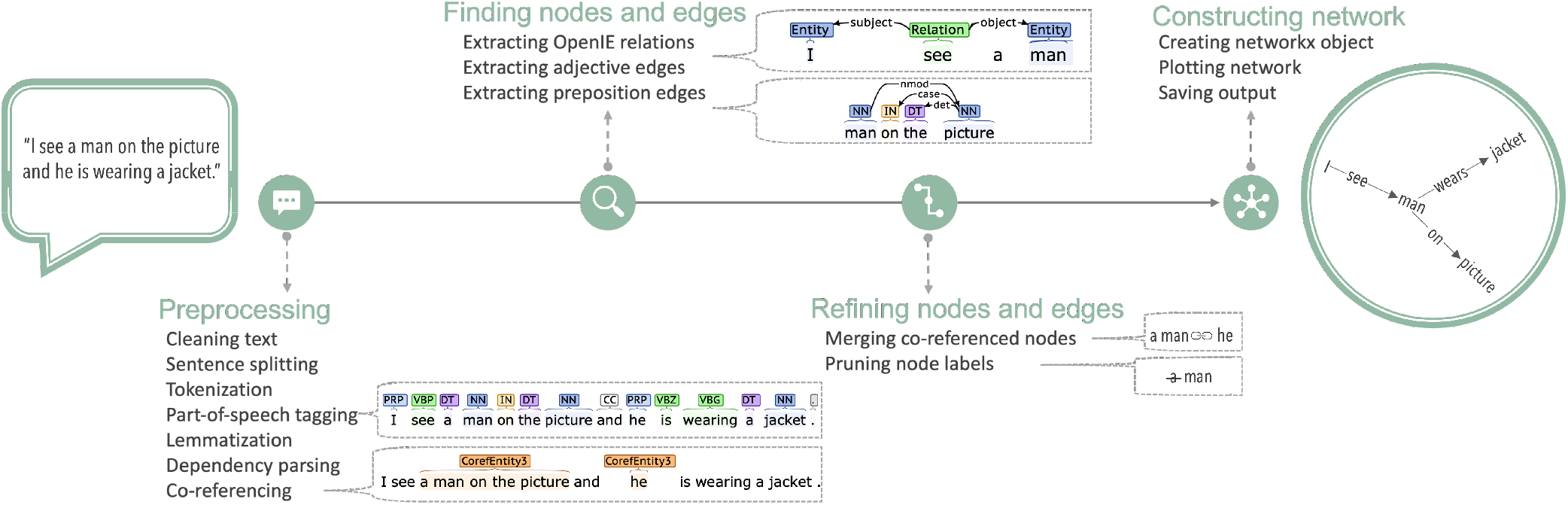
*Netts* processing pipeline. *Netts* takes as input a speech transcript and outputs a network representing the semantic content of the transcript: a *semantic speech network. Netts* combines modern, high performance NLP techniques to preprocess the speech transcript, find nodes and edge, refine these nodes and edges and construct the final semantic speech network.

#### Preprocessing

*Netts* first cleans the text by expanding common contractions (e.g. *I’m* becomes *I am*), and removing interjections (e.g. *Mh, Uhm*) and any transcription notes (e.g. timestamps). *Netts* then uses the CoreNLP library to perform sentence splitting, tokenization, part of speech tagging, lemmatization, dependency parsing and co-referencing on the transcript (18), using the default CoreNLP language model (for more details see Supplementary Note 1).

#### Finding nodes and edges

*Netts* submits each sentence to OpenIE5 to extract relationships between entities in the sentence using OpenIE5 default settings (19, 20). For example, from the sentence *I see a man* we identify the relation *see* between the entities *I* and *a man*. From these extracted relations, *netts* creates an initial list of network edges.

Next, *netts* uses the previously identified part of speech tags and dependency structure to extract additional edges defined by prepositions or adjectives: For instance, *a man on the picture* contains a preposition edge where the entity *a man* and *the picture* are linked by an edge labelled *on*. An example of an adjective edge is *dark background*, where *dark* and *background* are linked by an implicit *is*.

#### Refining nodes and edges

After creating the edge list, *netts* uses the previously identified co-referencing structure to merge nodes that refer to the same entity. For example, *a man* might be referred to by the pronoun *he*, or the synonym *the guy*. To ensure every entity is represented by a unique node, nodes referring to the same entity are merged by replacing them with the most representative node label (first mention of the entity that is a noun). In our example, *he* and *the guy* would be replaced by *a man*.

Node labels are then cleaned of superfluous words such as determiners, e.g. replacing *a man* with *man*.

#### Constructing network

Finally, *netts* constructs a semantic speech network from the edge list using the networkX library (21) and the network is plotted. The networkX object and the network image are saved for later analysis. The resulting graphs are directed and unweighted. An example semantic speech network is shown in Figure 2.

**Fig. 2.**
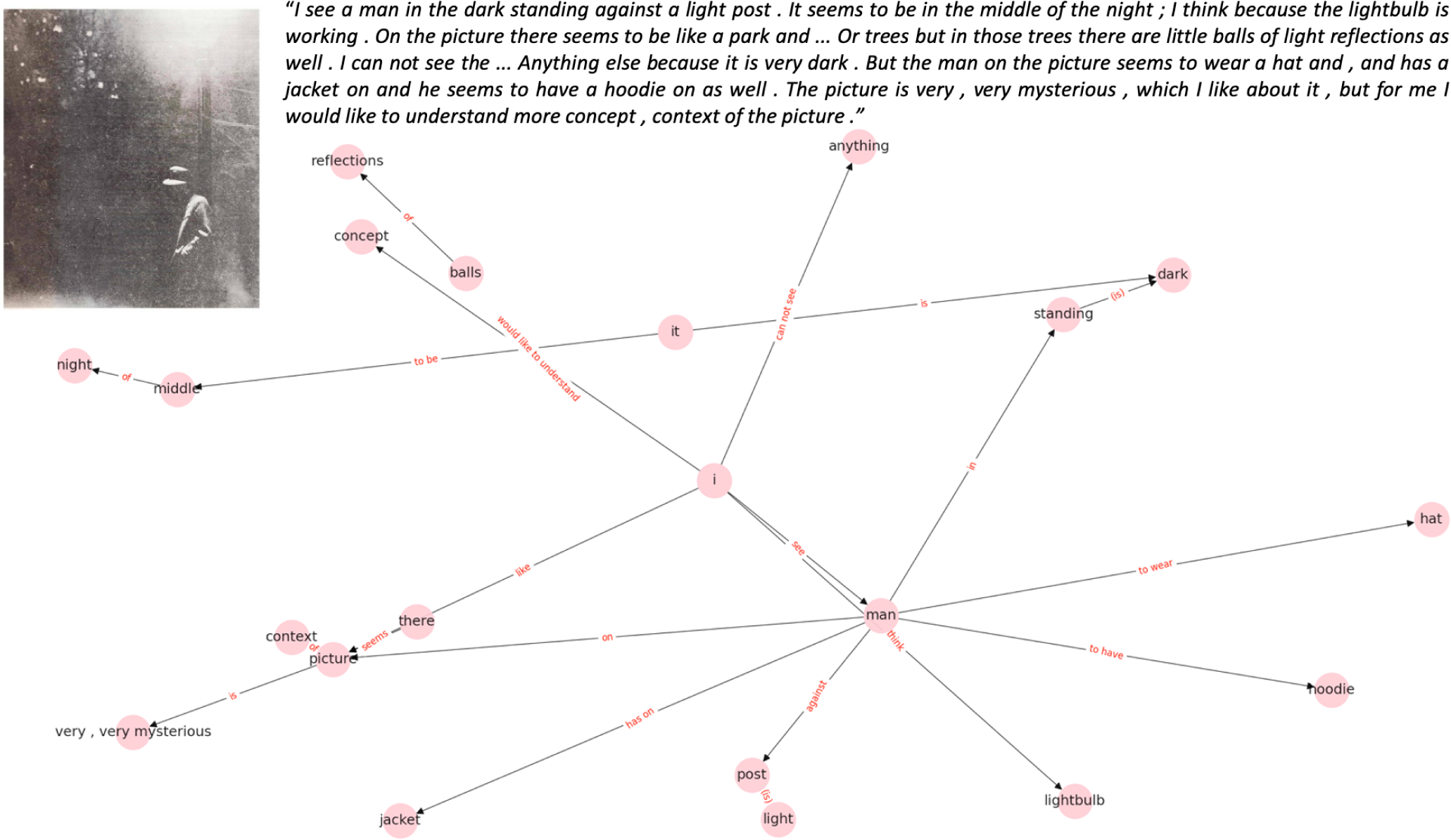
Example Speech Network. Semantic speech networks map the semantic content of transcribed speech. Nodes in the network represent entities mentioned by the speaker (e.g. *I, man*). Edges represent relations between nodes mentioned by the speaker (e.g. *see*). Top left figure inset shows the stimulus picture that the participant described. Top right figure inset is the speech transcript.

### Data

We applied *netts* to transcribed speech data from two studies: 1) a general population cohort (N=436), collected online, and 2) a clinical dataset including N=16 first episode psychosis (FEP) patients, N=13 healthy control subjects and N=24 subjects at clinical-high risk of psychosis (CHR-P), collected in person (total N=53) (22). CHR-P participants met the ultra-high risk criteria from the Comprehensive Assessment of At-Risk Mental States (CAARMS; (23)). Controls had no history of psychiatric illness. Further recruitment and demographic details are given in Supplementary Note 2. In both studies, participants described ambiguous pictures from the Thematic Apperception Test (TAT; (24)). Participants were presented with eight pictures from the TAT and instructed to talk about each picture for one minute while audio was recorded.

In the clinical dataset, participants additionally performed a story recall task, in which they were read six stories from the Discourse Comprehension Test (25) and asked to re-tell them with as many details as possible. In the following, we focus primarily on the picture description transcripts, which were available for both datasets, then assess whether we see similar clinical group differences in semantic networks with the story recall task.

The positive and negative syndrome scale (PANSS (26)) was used to measure symptom severity. The Thought and Language Index (TLI; (1)) was used to assess FTD.

The speech recordings were transcribed manually by a trained assessor, who was blinded to participant group status in the case of the clinical dataset.

All participants were fluent in English and gave written informed consent after receiving a complete description of the study. Ethical approval for the clinical study was obtained from the Institute of Psychiatry Research Ethics Committee, whilst ethical approval for the general population study was obtained from the King’s College London Research Ethics Committee.

### Speech Measures

#### Netts Measures

We constructed a semantic speech network for each transcript using *netts*. Overall, *netts* took ∼40 seconds processing time per 1 minute speech transcript. We then calculated the number of nodes and edges. We also calculated the number of connected components in each network, alongside their mean and median size. Here, a connected component is a subgraph of the original graph where all nodes are connected to each other by a path, ignoring edge directions. We decided to focus our analysis on connected components because when visualising networks from the general population dataset it was clear the networks often contained multiple connected components. Prior work on syntactic speech graphs has also shown size of connected components can be informative for psychosis (12).

All *netts* measures were compared to 1000 random Erdős-Rényi networks, matched for number of nodes and edges. The *netts* measures were then normalised to the random networks by z-scoring.

#### Additional NLP Measures

We compared our semantic speech graph measures to other NLP approaches that previously showed significant group differences in the clinical dataset (14). Specifically, we calculated semantic coherence, tangentiality, on-topic score, ambiguous pronoun count, and connectivity measures from the syntactic speech graphs proposed by (12).

#### Statistical Analysis

All measures were calculated for each transcript from each participant. To obtain a single value per participant, we calculated the mean average across the eight TAT picture descriptions.

We assessed normality of the measures using the Shapiro-Wilk test. Because some measures were not normally distributed and to mitigate the presence of potential outliers, we used the Mann-Whitney U-test to test for group differences (see Supplementary Table 1).

To explore the relationship between the *netts* measures and other speech measures, we calculated Pearson’s correlation coefficients between all measures. The resulting correlation matrix was clustered using the popular Louvain method for community detection (27).

Finally, we investigated the relationships between the *netts* measures and symptom and TLI scores, using linear regression, controlling for group membership as co-variates.

## Results

### General Public Networks

We first constructed semantic speech networks from the 2861 speech transcripts obtained from 436 members of the general public. 16 of the resulting networks were empty and therefore excluded from further analysis. Our final general public sample therefore consisted of 2845 networks. The resulting semantic speech networks consisted of 15.77 ± 5.03 nodes and 15.04 ± 5.61 edges on average; see Figure 3.

**Fig. 3.**
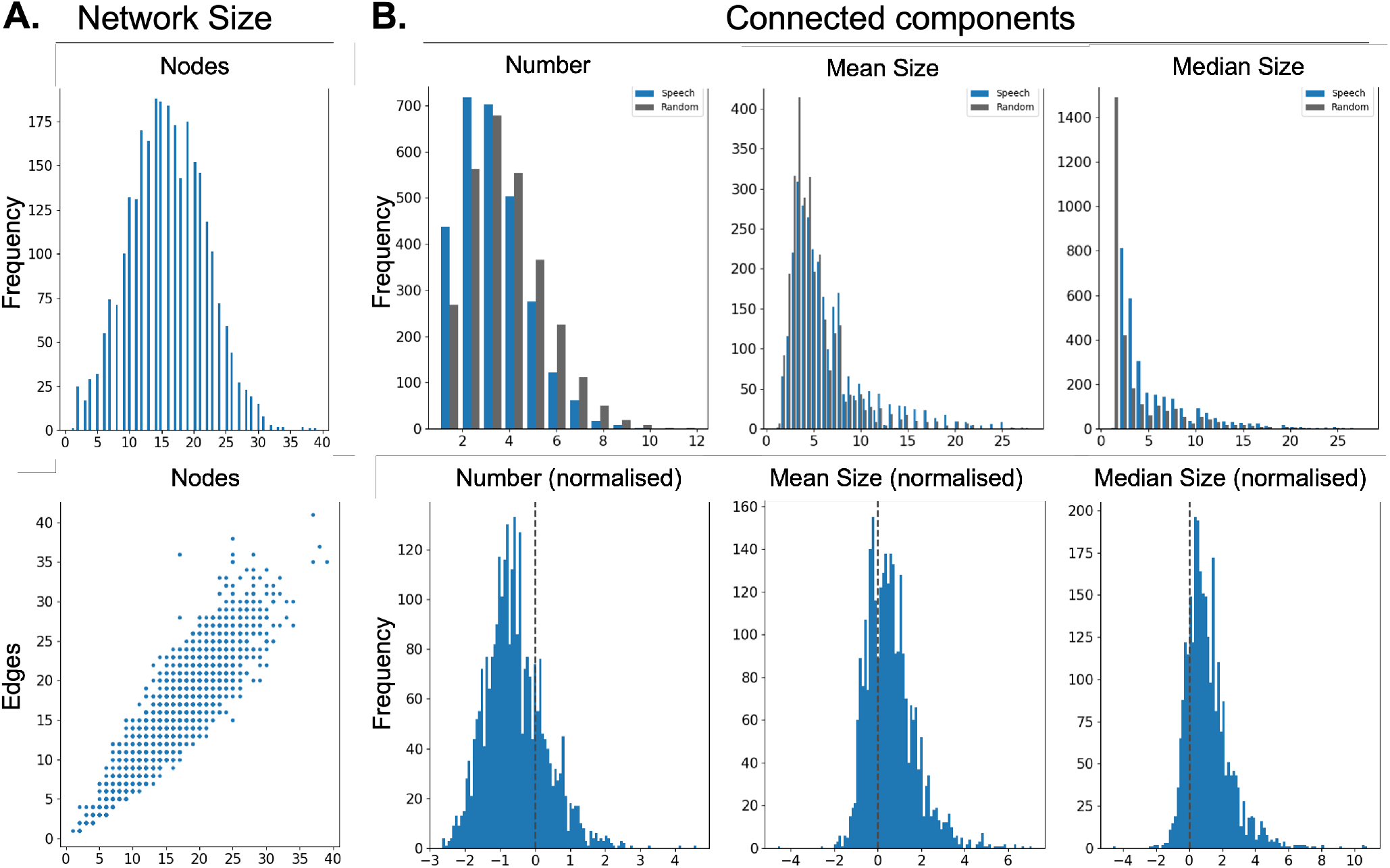
General Public Networks. Semantic speech networks differ in their properties from random networks. **A**. Histogram for number of nodes and scatter plot showing the relationship between number of nodes and number of edges of semantic speech networks from the general public. **B**. Top row shows number, mean size and median size of the connected components in the speech graphs (blue bars) and a randomly chosen subset of the size-matched random graphs (grey bars). Bottom row shows normalised number, mean size and median size of the connected components in speech graphs.

#### Speech networks had fewer and larger components than random networks

Compared to size-matched random networks, the speech networks had fewer connected components (t = -28.51, p = < 0.00001, Mean ± Std: -0.55 ± 0.41); see Figure 3. These connected components were also significantly larger than the connected components of size-matched random networks (Mean: t = 23.19, p = < 0.00001; Median: t = 35.0, p = < 0.00001; Figure 3).

### Networks in the clinical dataset

The clinical dataset consisted of 415 transcripts. FEP patients spoke trendwise fewer words and more sentences than controls, resulting in shorter sentences (words: z = 1.73, p = 0.08; sentences: z = -2.39, p = 0.02, mean sentence length: z = 2.83, p < 0.01).

There was no difference in the number of words or sentences between CHR-P participants and either FEP patients or controls (all p > 0.12).

#### Patient networks were smaller

Differences in amount of speech were reflected in the semantic speech networks. FEP networks had fewer nodes (z = 2.33, p = 0.02) and fewer edges (z = 1.97, p = 0.048) than healthy control networks. Numbers of nodes and edges did not differ between CHR-P participants and either FEP patients or controls (all p > 0.13). Representative FEP patient and control networks are shown in Figures 4D and 4E.

**Fig. 4.**
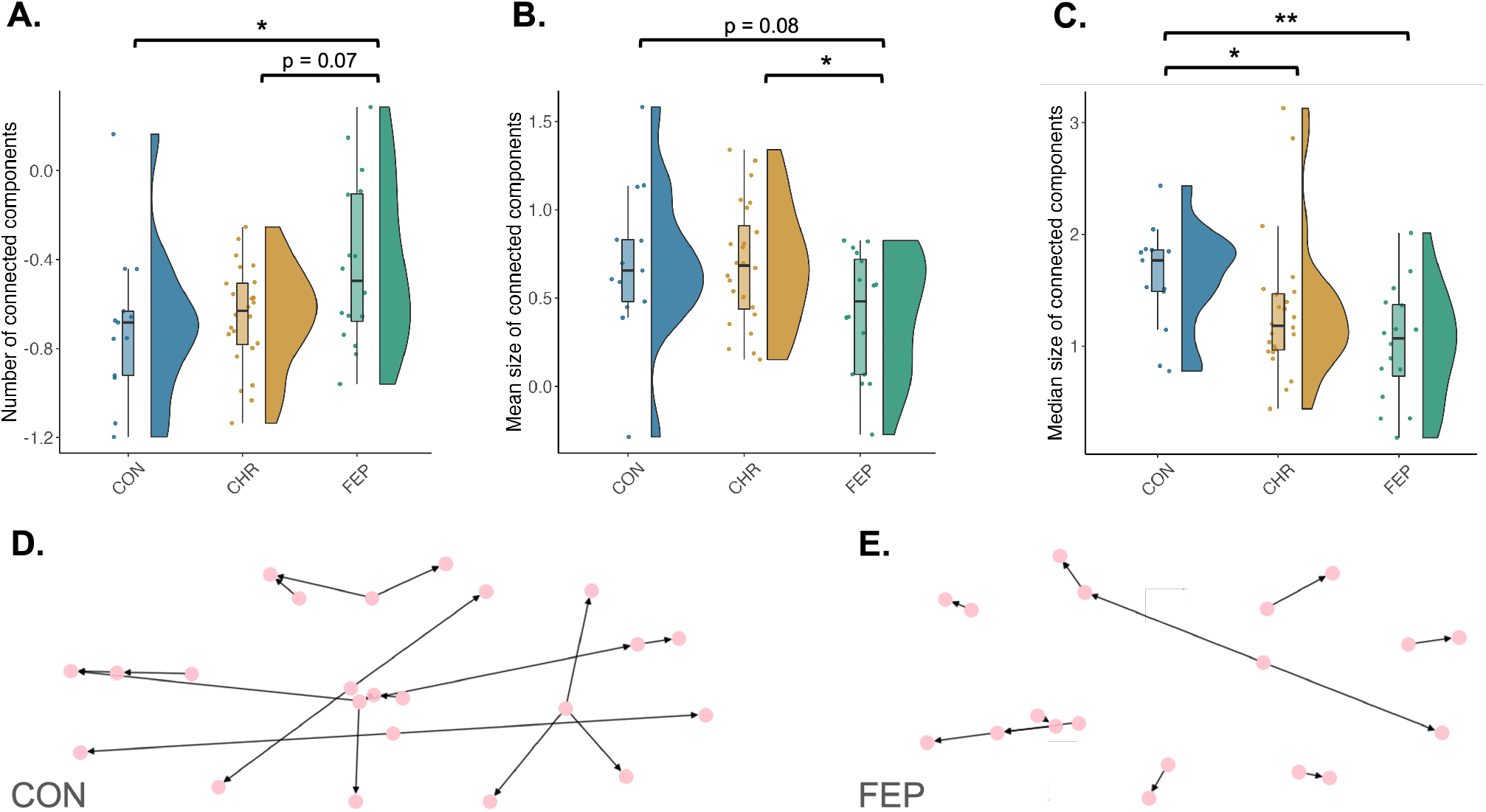
Clinical Networks differ between groups. A) Number of connected components, B) mean connected component size and C) median connected component size showed differences between the FEP patient (FEP), clinical high risk (CHR-P) and healthy control groups (CON). Network measures shown are normalised to random networks. *indicates significant p-values at p < 0.05. **indicates significant p-values at p < 0.01 **D and E**. Semantic speech networks of patients had more and smaller connected components than the networks of healthy controls. **D** shows a typical network from a healthy control participant and **E** shows a typical network from a first episode psychosis patient. Plots A-C were produced using the Raincloud package (28) and D-E using networkX (21).

#### Patient networks had more and smaller components

The normalised number of connected components was higher for FEP patient networks than healthy control networks (z = -2.00, p = 0.046; Figure 4A). FEP patients also had a trend-wise higher number of connected components than CHR-P participants (z = 1.81, p = 0.07). There was no significant difference in the normalised number of connected components between CHR-P participants and controls (z = -0.79, p = 0.43)).

Mean connected component size was trendwise smaller for FEP patients than healthy controls (z = 1.73, p = 0.08; Figure 4B) and smaller for FEP patients than CHR-P participants (z = -2.01, p = 0.04). There was no significant difference in mean connected component size between CHR-P participants and healthy controls (z = 0.13, p = 0.90).

We also calculated the median connected component size, which should be less influenced than mean size by the one or two very large connected components speech networks often contain. Both FEP patients and CHR-P participants had significantly smaller median connected component size compared to controls (z = 2.70, p < 0.01 for FEPs, z = 2.14, p = 0.03 for CHR-P subjects; Figure 4C). There was no difference in median connected component size between CHR-P participants and FEP patients (z = -1.16, p = 0.25).

#### Sensitivity analyses

Our normalisation procedure controls for network size. However, we performed an additional sensitivity analysis to test whether the group differences in semantic speech network measures remained significant when controlling for number of words. To that end, we constructed a linear model of the network measure as a function of group and included number of words as a covariate.

The FEP patient-control difference in number of connected components remained significant after controlling for number of words (Significant overall regression model: F(3, 48) = 3.33, p = 0.03, with significant effect of patient: beta = 0.24, p = 0.04), as did the FEP patient-control difference in the median size of connected components (Significant overall regression model: F(3, 48) = 4.868, p < 0.01, with significant effect of patient: beta = -0.51, p = 0.01). The difference in median size of connected components between CHR-P participants and healthy controls did not survive controlling for number of words (no significant effect of CHR-P participant: beta = -0.29, p = 0.12). There was no significant group difference between FEP, CON and CHR-P networks in mean size of connected components after controlling for number of words (F(3, 48) = 1.97, p = 0.13).

#### Semantic speech networks may capture novel information

Figure 5 shows a heatmap of the correlations between the semantic speech network measures and other NLP measures, calculated using the clinical dataset. The black lines denote communities detected using the Louvain method (27).

**Fig. 5.**
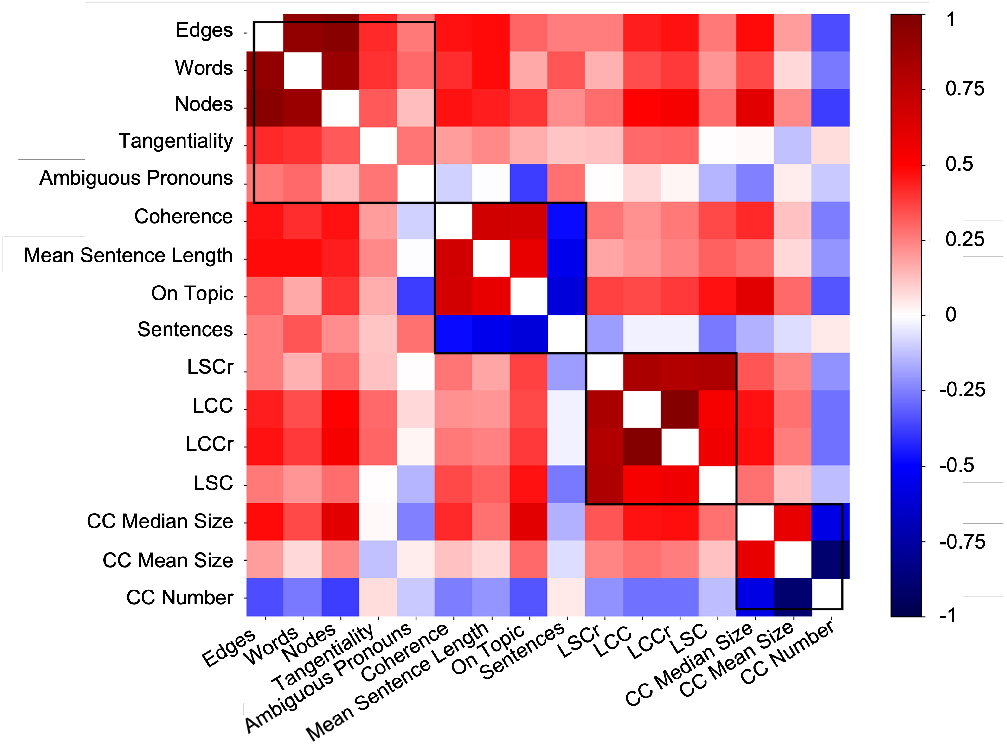
Clustered speech measures. Semantic speech network measures captured signal complementary to other NLP measures. Shown is a heatmap of Pearson’s correlations between semantic speech network measures and NLP measures in the clinical dataset. Black lines indicate communities detected using the Louvain method. The measures used in this analysis were the novel netts measures, as well as basic transcript measures and established NLP measures. Netts measures were number of connected components (CC Number), mean connected component size (CC Mean Size) and median connected component size (CC Median Size). Basic transcript measures were number of words, number of sentences and mean sentence length. Established NLP measures included Tangentiality, Ambiguous Pronouns, Semantic Coherence (Coherence), On-Topic Score (On Topic) taken from (14). Additionally, syntactic network measures were taken from (14) and included number of nodes in the largest strongly connected component of syntactic networks (LSC), number of nodes in the largest weakly connected component of syntactic networks (LCC), as well as the LSC and LCC normalised to random networks (LSCr, LCCr) (12)

As expected, the number of nodes and number of edges of the semantic speech networks clustered with the number of words in the transcript. The connected component measures (CC Number, CC Median Size, CC Mean Size) formed their own community, and did not cluster with number of words, number of sentences, measures of network size, or any other NLP measures.

#### Component size was associated with TLI scores

Supplementary table 2 shows the relationships of the network measures with the PANSS symptom scores and the TLI. Median connected component size was negatively related to TLI Negative scores (t = -2.02, p = 0.049, beta = -0.32). The number of connected components and the mean size of connected components predicted PANSS Negative scores (CC Number: t = -2.55, p = 0.02, beta = 5.76; CC Mean Size: t = -2.12, p = 0.046, beta = -4.24), with participants scoring higher on the PANSS Negative scale having more and smaller connected components. However, none of these relationships survived Bonferroni-correction (significance threshold corrected for 6 × 3 comparisons: p = 0.0028).

Inspecting the TLI subscales, we observed a relationship between median connected component size and poverty of speech (t = -2.33, p = 0.02, beta = -0.329678; Supplementary Table 3). Number of connected components and mean connected component size were related to peculiar logic (CC Number: t = 2.6, p = 0.01, beta = 0.216731 ; CC Mean Size: t = -2.09, p = 0.04, beta = -0.148592). However, none of these relationships survived Bonferroni-correction (significance threshold corrected for 7 × 3 comparisons: p = 0.0024). After poverty of speech, median component size was most strongly associated with looseness of associations, although not significantly (t = -1.45, p = 0.15; beta = -0.2).

#### Story networks show FEP patient-control differences in size, but not in connected components

Finally, we assessed task dependency of our results using speech generated from a story recall task.

Although number of words and number of sentences did not differ between FEP patients and healthy controls (words: z = 1.46, p = 0.15, sentences: -1.18, p = 0.24), FEP patients showed reduced sentence length (z = 2.56, p = 0.01). FEP patients also showed reduced mean sentence length compared to CHR-P participants (z = -2.58, p < 0.01). FEP patients spoke fewer words than CHR-P participants (z = -2.16, p = 0.03), but did not differ in number of sentences (z = 0.7, p = 0.49). There was no difference in the number of words and sentences or mean sentence length between CHR-P participants and controls (all p > 0.37).

This difference between FEP patients and CHR-P participants in the amount of speech was reflected in the semantic speech networks. FEP patient networks consisted of fewer nodes (z = -2.44, p = 0.01) and fewer edges (z = -2.9, p < 0.01) than CHR-P networks. Network size did not differ between healthy controls and either FEP patients or CHR-P participants (all p > 0.1), although there was a trendwise difference in the number of edges between controls and FEP patients (z = 1.87, p = 0.06).

The normalised number of connected components did not differ between groups (all p > 0.18). We also found no significant group differences in the mean and median size of the connected components (all p > 0.25).

Finally, in a post-hoc analysis to compare the characteristics of the story recall and picture description networks, we calculated the ratio of edges to nodes for all networks. The semantic networks from the story re-telling task had significantly more edges per node than those from the picture description task (t = -11.87, p < 0.0001).

## Discussion

We developed a novel algorithm, and software package, “*netts*” to map the semantic content of a speech transcript as a network, and investigate the nature of semantic speech networks in psychosis.

*Netts* maps semantic networks by 1) identifying the entities described in a speech transcript (the nodes), 2) extracting the semantic relationships between entities (the edges) and plotting the resulting graph. This approach has several advantages. First, relationships between entities can be extracted from relatively distant parts of the transcript. The tool is also computationally fast, and robust against artefacts typical for transcribed speech, such as interjections (Um, Ah, Err) and word repetitions (I think the the man). The algorithm therefore lends itself to the automated construction of speech networks from large datasets.

Having developed the tool, we initially applied it to speech transcripts from the general population. The networks had fewer and larger connected components than size-matched random Erdo?s-Rényi networks. This distinction from random networks reflects the nature of semantic speech networks, and the fact that when describing the picture people tend to link different aspects of the picture together in a way that is intrinsically non-random.

In our clinical sample, semantic speech networks from FEP patients had fewer nodes and edges than those from healthy controls for the picture description, but only trendwise differences in edges for the story recall task. CHR-P subjects had more nodes and edges than FEP patients for the story recall task, but not the picture description task. These observations are in-line with the group differences in the number of words spoken, and poverty of speech in psychosis.

We also explored whether there were group differences in the number and size of the networks’ connected components, motivated by the loosening of associations often observed in FEP. In the picture description task, FEP patient networks were more fragmented than control networks, with more, smaller connected components.

Relationships between the network connectivity measures and the TLI and PANSS scores were strongest for the negative TLI and negative PANSS scores, but did not survive correction for multiple comparisons. We note that a large majority of participants scored zero for TLI Negative (71%), peculiar logic (86%) and poverty of speech (75%), suggesting a flooring effect could be obscuring significant relationships. Interestingly, after poverty of speech, median component size was most strongly associated with looseness of associations. This relationship did not reach significance, though here again a flooring effect might be obscuring a significant relationship because 80% of participants in the clinical dataset scored zero for looseness of associations.

The increased fragmentation in FEP patient networks compared to controls was not observed with the story recall task. This could be a result of the differing cognitive demands of the two tasks and the probing of distinct mental processes (working memory vs requiring participants to spontaneously connect entities with each other). We note that semantic speech graphs from the story recall task also had more edges per node than those from the picture description task, again likely reflecting the more structured nature of the recalled stories compared to spontaneous speech. Further work is required to fully understand these task dependencies. Nonetheless, our results again suggest the need for careful consideration of the choice of tasks to elicit speech responses from participants (12, 14).

As expected, in a clustering analysis, the number of nodes and edges in the semantic speech networks clustered with the number of words. The number and size of connected components formed their own community, independent of previously reported NLP measures (14) including syntactic graph measures (11) and semantic coherence. The connected components in semantic speech networks might therefore capture signal beyond information contained in established NLP measures or relating to the amount of speech.

Overall, *netts* allows us to map the semantic content of speech as a network, opening the door to more detailed analyses of the semantic content of speech, and how this is altered in FTD. Measures of network size and connectivity showed significant group differences between FEP patients, CHR-P subjects and healthy controls and provided complementary information to established NLP measures. We hope that the open availability of *netts* will allow other researchers to explore this new perspective on how disorganized speech is manifest in psychosis.

### Limitations

We examined relatively basic topological properties of semantic speech networks, e.g. size and number of connected components. Nonetheless, semantic speech networks also include additional information. For example, *netts* records temporal information about the order in which edges were formed in the networks, which could be used to give an indication of network coherence or staying on topic. The words associated with nodes and edges could also be represented as vectors using word embedding methods, to provide rich node labels. These more advanced network features could provide additional power for deep phenotyping of psychotic disorders.

*Netts* is currently only available for the English language and would need to be adapted for use with other languages.

Finally, the modest sample size of this study, though similar to those used in previous studies is unlikely to fully capture the diverse phenotype of psychosis (29) and the heterogeneity of CHR-P subjects. It is therefore crucial for future studies to test the generalisability of these results. To support such efforts, we will release *netts* as a free and open-source Python package on publication of this paper. For peer-review purposes, a pre-release of the toolbox is available at: https://pypi.org/project/netts/0.2.0rc1/.

## Data Availability

All data produced in the present study are available upon reasonable request to the authors.

## Author Contributions

CRN wrote the network algorithm, packaged and documented the *netts* toolbox, conceived, designed and performed the analysis, and wrote the original draft of the manuscript. OG, IS and HD packaged and documented the *netts* toolbox. NC contributed to the NLP analysis. PEV contributed to the network analysis. KD, JO, TG-D, TJS and PM collected the data. KD and TJS acquired the funding for the general population data collection and conceived the population cohort study design. PM acquired the funding for the clinical data collection and conceived the original clinical study design. SEM conceived and designed the network algorithm, conceived and performed the analysis, and acquired the funding for the development of the network algorithm and toolbox. All authors discussed the data interpretation and reviewed and revised the manuscript.

## Acknowledgments

We thank the services users and volunteers who took part in this study, and the members of the Outreach and Support in South London (OASIS) team who were involved in the recruitment, management and clinical follow-up of the participants reported in this manuscript. SEM was supported by the Accelerate Programme for Scientific Discovery, funded by Schmidt Futures and a Fellowship from The Alan Turing Institute, London. PEV is supported by a fellowship from MQ: Transforming Mental Health (MQF17_24). This work was supported by The Alan Turing Institute under the EPSRC grant EP/N510129/1, the UK Medical Research Council (MRC, grant number G0700995), the NIHR Cambridge Biomedical Research Centre (BRC-1215-20014), the King’s Health Partners (Research and Development Challenge Fund) and the National Institute for Health Research (NIHR) Mental Health Biomedical Research Centre at South London and Maudsley NHS Foundation Trust and King’s College London. The views expressed are those of the author(s) and not necessarily those of the NHS, the NIHR, MRC or the Department of Health and Social Care. The funder had no influence on the design of the study or interpretation of the results.

## Supplementary Note 1: Details on *netts* preprocessing

*Netts* preprocesses the speech transcript by expanding common English contractions, removing interjections and removing any transcription notes. These steps are customizable, e.g. with the option to pass a user-defined text file of transcription notes that should be ignored by *netts*.

*Netts* then uses CoreNLP to perform sentence splitting, tokenization, part of speech tagging, lemmatization, dependency parsing and co-referencing on the transcript (18) with the default language model implemented in CoreNLP (Universal Dependencies English Web Treebank model (30)). These NLP techniques are briefly described in the following. The transcript is first split into sentences (sentence splitting) and further split into meaningful entities, usually words (tokenization). Each word is then assigned a part of speech label, indicating whether it is a verb, noun, or another part of speech (part of speech tagging). Each word is also assigned their dictionary form or lemma (lemmatization) and the grammatical relationship between words is identified (dependency parsing). Finally, any occurrences where two or more expressions in the transcript refer to the same entity are identified (co-referencing), for example where a noun *man* and a pronoun *he* refer to the same person.

## Supplementary Note 2: Details on general public dataset demographics

Participants were recruited from the general public as part of an online study through online advertisement (N=444).Recruitment took place over a seven-day period via the online recruitment platform Prolific (https://prolific.co), which enabled participant anonymity. Requirements for participant inclusion included: age 16-40yrs, fluency in the English language, and residence in the UK. Participants were compensated for their time at a rate of £5.00 per hour. 48 transcripts were excluded from the analysis due to poor audio quality, resulting in 8 participants being fully excluded from the sample and a final dataset of N=436 participants. Participants (120 male) were on average 28.03 ± 6.27 years old (range 18 - 40 years), and included 82% English native language speakers. 12% of participants self-reported a current mental health diagnosis. Demographic data was unavailable for 22 participants.

This was a cross-sectional population study. Participants were provided a webpage link through Prolific to an online testing platform Gorilla (https://gorilla.sc/). Individuals were given information about the study and provided their informed consent on Gorilla. Participants could use their own internet-accessible devices including smartphones, tablets, or computers. Ethical approval was obtained through King’s College London [REMAS MRA-19/20-19444]. Data collected via Gorilla was compliant with EU GDPR2016 and Gorilla’s ‘Data Processing Agreement’, consistent with NIHR guidance.

Participants completed three learning and decision making online tasks and the speech task as described previously (Spencer et al., 2021). Only speech data are reported here.

### A. Clinical Dataset

Participants were recruited as described in (22). Briefly, FEP patients were recruited from the South London and Maudsley NHS Foundation Trust. CHR-P participants were recruited from the Outreach and Support in South London (OASIS) service, which is part of the Pan-London Network for Psychosis-prevention (22). CHR-P participants met ultra-high risk criteria assessed with the Comprehensive Assessment of At-Risk Mental States (CAARMS; (22)). Healthy control participants had no previous or current history of psychiatric illness and no family history of psychosis and were recruited from the same geographical area by local advertisement and through contacts of CHR-P participants after receiving written permission. Healthy controls were matched to the CHR-P and FEP participants for age and gender. Demographics for the three groups have been reported elsewhere (22) (14).

If the participant stopped talking during the minute they were prompted to continue by the interviewer.

Participants were excluded where a history of a neurological or medical disorder, history of head injury, or alcohol or illicit substance misuse or dependence was present.

For the clinical dataset the recording of one participant was excluded due to poor audio quality, leaving 52 participants. One other participant had only seven picture descriptions, but was still included in the analysis.

## Supplementary Note 3: Normality

**Table 1.** Normality of network and NLP measures. Normality of network measures and NLP measures calculated from picture description (TAT) transcripts was assessed using a Shapiro-Wilk test. Tests were performed for each group of the clinical dataset separately. Shown are p-values of the Shapiro-Wilk test. *indicates significant p-values at p < 0.05.

| Measure | Group |  |  |
| --- | --- | --- | --- |
|  | CON | CHR-P | FEP |
| Nodes | 0.32 | 0.82 | 0.84 |
| Edges | 0.69 | 0.89 | 0.49 |
| CC Number | 0.1 | 0.8 | 0.23 |
| CC Mean Size | 0.73 | 0.66 | 0.11 |
| CC Median Size | 0.94 | **0.002\*** | 0.93 |
| Words | 0.67 | 0.76 | 0.99 |
| Sentences | 0.6 | **0.02\*** | 0.17 |
| Mean Sentence Length | **0.00009\*** | 0.38 | 0.26 |
| Coherence | 0.74 | 0.36 | 0.21 |
| Tangentiality | 0.12 | 0.9 | 0.21 |
| On-topic | 0.91 | 0.33 | 0.69 |
| LCC | 0.4 | 0.23 | 0.16 |
| LSC | 0.73 | 0.16 | **0.03\*** |
| LCCr | 0.1 | 0.81 | **0.04\*** |
| LSCr | **0.03\*** | 0.78 | 0.45 |

## Supplementary Note 4: Semantic speech network measure associations with symptom / TLI scores

**Table 2.** Relationship between symptoms and network measures. Symptoms were measured using the Thought and Language Index (TLI) and the Positive And Negative Syndrome Scale (PANSS). Relationships were assessed using a linear regression of symptom measures controlling for group membership as a covariate. Results are shown as t-statistics with p-values in brackets for the network measures as a predictor of symptom score. *indicates significance before correcting for multiple comparisons. None of the relationships survived Bonferroni-correction (significance threshold corrected for 6 × 3 comparisons: p = 0.0028).

| Symptoms / TLI scores | Connected components in semantic speech networks |  |  |
| --- | --- | --- | --- |
|  | Number | Mean size | Median size |
| TLI Total | 0.98 (0.33) | -0.95 (0.35) | -1.26 (0.21) |
| TLI Positive | 0.54 (0.59) | -0.61 (0.55) | -0.56 (0.58) |
| TLI Negative | 0.85 (0.34) | -0.58 (0.57) | -2.02 (0.049)* |
| PANSS General | 0.51 (0.61) | -0.76 (0.46) | -0.8 (0.43) |
| PANSS Positive | 0.92 (0.37) | -0.95 (0.35) | -0.43 (0.67) |
| PANSS Negative | 2.55 (0.02)* | -2.12 (0.046)* | -1.38 (0.18) |

## Supplementary Note 5: Semantic speech network measure associations with TLI subscales

**Table 3.**
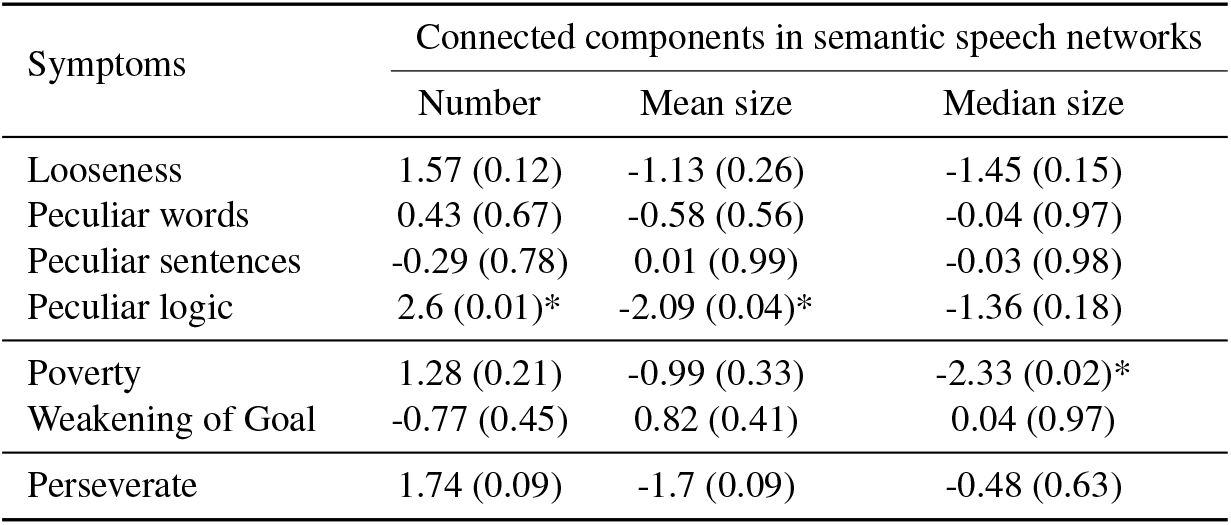
Relationship between TLI subscales and network measures. Relationships were assessed using a linear regression of symptom measures controlling for group membership as a covariate. Results are shown as t-statistics with p-values in brackets for the netwrok measures as a predictor of TLI subscale. *indicates significance before correcting for multiple comparisons. None of the relationships survived Bonferroni-correction (significance threshold corrected for 7 × 3 comparisons: p < 0.01).

